# Identification of novel nutrient-sensitive gene regulatory networks in amniocytes from fetuses with spina bifida

**DOI:** 10.1101/2022.03.29.22273067

**Authors:** Marina White, Jayden Arif-Pardy, Kristin L Connor

## Abstract

Neural tube defects (NTDs) remain among the most common congenital anomalies. Contributing risk factors include genetics and nutrient deficiencies, however, a comprehensive assessment of nutrient-gene interactions in NTDs is lacking. We applied a nutrient-focused gene expression analysis pipeline to identify nutrient-sensitive gene regulatory networks in amniocyte gene expression data (GSE4182) from fetuses with NTDs (cases; n=3) and fetuses with no congenital anomalies (controls; n=5). Differentially expressed genes (DEGs) were screened for having nutrient cofactors. Nutrient-dependent transcriptional regulators (TRs) that regulated DEGs, and nutrient-sensitive miRNAs with a previous link to NTDs, were identified. Of the 880 DEGs in cases, 10% had at least one nutrient cofactor. DEG regulatory network analysis revealed that 39% and 52% of DEGs in cases were regulated by 22 nutrient-sensitive miRNAs and 10 nutrient-dependent TRs, respectively. Zinc- and B vitamin-dependent gene regulatory networks (Zinc: 10 TRs targeting 50.6% of DEGs; B vitamins: 4 TRs targeting 37.7% of DEGs, 9 miRNAs targeting 17.6% of DEGs) were dysregulated in cases. We identified novel, nutrient-sensitive gene regulatory networks not previously linked to NTDs, which may indicate new targets to explore for NTD prevention or to optimise fetal development.

## Introduction

Neural tube defects (NTDs) are among the most prevalent congenital anomalies^1^. Complex genetic and environmental contributions underly around 75% of NTDs, which have multifactorial causation^2^ and can associate with multiple comorbidities, including placental maldevelopment and function^3^, fetal growth restriction, and preterm birth^3,4^. Folate and vitamin B12 have a well-recognised contribution to the etiology of isolated NTDs^5^, while other nutritional factors, including inositol^6^, vitamin D^7^, and choline^8^ and essential trace elements^9^, may also contribute but are less well-studied. Accordingly, widespread and targeted nutritional prevention strategies, such as folic acid fortification and periconceptional supplementation, have decreased NTD incidence^10^. However, that more than 300,000 infants remain affected by NTDs annually^1^ highlights the importance of improving knowledge on their causation to inform further strategies for prevention.

The role of genetics in NTD etiology is supported by associations between NTDs and chromosomal anomalies, heritable susceptibility to NTDs^11^, and the identification of the C677T polymorphism in the methylenetetrahydrofolate reductase (*MTHFR*) gene as a genetic risk factor for NTDs^12^. Distinct genetic signatures that associate with NTDs have also been revealed through large-scale genomic analysis^13,14^, which can provide an integrated, global view of the mechanisms regulating normal and pathological development and aid in the identification of biomarkers for prevention and diagnosis^15^. Some research on gene expression signatures in NTDs has been performed using amnioctyes^13,14^, which are a heterogeneous population of fetal cells in the amniotic fluid^16^. Amniocytes comprise pluripotent, lineage-committed, and mature differentiated cells, and are mostly derived from embryonic/fetal tissues^16,17^. Amniocyte transcriptome profiling has detected the expression of genes with known tissue-specific expression in fetal organs, including the brain and placenta^16^, suggesting that amniocytes may be uniquely positioned to inform on gene expression signatures in NTDs, and potentially placental-related comorbidities. While transcriptome profiling can help provide a gene-, pathway-, and network-level understanding of the gene expression signatures that associate with NTDs, there is also a need to integrate knowledge on gene-environment, including nutrient, interactions to work towards a more complete understanding of NTD phenotypes, and to identify target candidates for prevention.

Adequate nutritional resources are essential for ensuring normal embryonic and fetoplacental growth and development. Nutrients play important roles in DNA stability and repair and gene expression regulation, and host genetics influence nutrient metabolism and bioavailability^18^. Research to-date has largely focused on the contribution of genetic variants in the folate pathway to NTD risk^11^. However, knowledge is limited on whether, and the extent to which, *other* nutrients and their interactions with genes can explain NTD etiology. This is a critical gap to fill, given that fetal NTDs persist even in countries with folic acid food fortification^19^, and in pregnant people with normal blood folate levels^20^. Comorbidities associated with NTDs, including altered placental phenotype^3^, poor fetal growth^4^, and early birth^3,4^, may also be responsive to, or driven by, altered nutrient bioavailability^21^. However, to date, no study has integrated transcriptome network analysis with field knowledge on nutrient-gene interactions in the context of NTDs or their comorbidities.

Here, we applied an untargeted, nutrient-focused gene expression analysis pipeline to help address knowledge gaps and generate new hypotheses on the role of nutrient-gene interactions in NTDs. Using data from a small cohort study, we first aimed to examine the degree to which differentially expressed genes (DEGs) in fetuses with spina bifida (cases) were micronutrient dependent. We next aimed to discover novel nutrient-sensitive gene regulatory networks dysregulated in NTDs. We hypothesised that multiple nutrient-gene interactions would be evident in gene signatures associated with NTDs, and that clear nutrient-sensitive miRNA and TR ‘regulons’ (a group of genes belonging to a common regulatory network) would emerge among DEGs.

## Methods

### Data source and pre-processing

Amniocyte gene expression data for fetuses with spina bifida (cases; n=4) and fetuses with no congenital anomalies (controls; n=5) were obtained from Gene Expression Omnibus (GEO; GSE4182) on June 28, 2021, and re-analysed. Secondary analysis of anonymized data is exempt from ethics approval by the Carleton University Research Ethics Board. As part of a case-control study in Budapest, Hungary, amniocytes were isolated from amniotic fluid samples collected during amniocentesis and stored at -80°C until RNA extractions were performed^14^. Amniocyte transcriptome sequencing was carried out using the Affymetrix Human Genome U133 Plus 2.0 Array, and microarray deposited in GEO in November 2006^14,22^. Sample characteristics are provided in Supplementary Table S1. All three cases had a spina bifida diagnoses, and one case also had anencephaly. Gestational age at time of amniotic fluid collection ranged from 13-19 weeks for cases and 17-19 weeks and 5 days for the control group. Each control sample was pooled from two patient amniotic fluid samples from pregnancies undergoing routine amniocentesis because of a maternal age >35 years, and thus included amniotic fluid from 10 controls total.

All data pre-processing were performed in R software (v3.6.1) using the Bioconductor (v3.12) Affy package^23^. Raw microarray data (.CEL files) were downloaded and normalised using the robust multi-array average method^24^. Expression values were visualised using box-and-whisker plots and outliers were identified using sample clustering and uniform manifold approximation and projection (UMAP)^25^. Where multiple probesets corresponded to a single gene, a representative probeset was chosen for each gene, based on recommended methods (Supplementary Table S2)^26^. Probeset identifiers were annotated with Entrez gene IDs^27^.

### Differential gene expression analysis

DEGs between cases and controls were identified (eBayes; R [v3.6.1])^28^. Statistical significance was determined after false discovery rate correction at *q* value <0.05 and an absolute log_2_ fold change (FC)≥2^29^. Functional analysis of the DEG list was performed using the Protein ANalysis THrough Evolutionary Relationships (PANTHER) classification system (v16.0)^30^.

### Identifying nutrient cofactors of differentially expressed genes

To determine which of the DEGs coded for proteins that were micronutrient dependent (i.e., they coded for proteins with one or more micronutrient [vitamin or mineral] cofactor), we created a DEG-nutrient interaction network. Using a previously published dataset of protein/gene-nutrient (cofactor) interactions^31^, we identified DEGs coding for a cofactor-interacting protein and their associated cofactors. This dataset on protein-nutrient interaction networks integrates knowledge on nutrient-protein (gene) interactions from the Metal MACiE25, Uniprot23, Expasy24 and EBI CoFactor databases. Applying a systems-approach to nutrient-gene interaction analysis is key for understanding the ways in which multiple micronutrients, their metabolic products, and interacting proteins function in biological processes, and for obtaining a more complete view of nutritional contributions to health and disease^31^. The PANTHER classification system (version 16.0) was then used to functionally annotate DEGs that were nutrient-dependent^30^, and we visualised DEG-nutrient relations as a network using Cytoscape (version 3.8.2)^32^.

### Nutrient-sensitive miRNA targetome analysis

Next, to construct nutrient-sensitive miRNA regulatory networks for DEGs in cases, we first curated a list of nutrients that have known relevance to NTDs. This included cofactors for one carbon metabolism and nucleotide synthesis (choline^33^, vitamins B1^34-36^, B6^33,35,36^, B9 [folate]^35,36^, and B12^33,35^), as well as inositol^37^, iron^36^, and vitamin D^7^. We next used miRWalk (version 2.0), a platform that integrates data on validated and predicted miRNA-target (gene) interactions, to identify miRNAs that are known to interact with biological processes related to these nutrients, or with activity known to be sensitive to these nutrients^38^. We then cross-referenced the list of miRNAs sensitive to, or associated with, these NTD-related nutrients against a curated inventory of miRNAs that have been associated with NTDs or neural tube closure in previous animal and human studies, which was also built using miRWalk and through manual searching in PubMed (search string: “neural tube*” AND “miRNA”). Targetome files with miRNA-target gene network data (miRNA targetomes), limited to experimentally validated miRNA-target interactions (miRTarBase)^38,39^, were then retrieved for the miRNAs that were found in both lists (i.e., miRNAs whose activity was sensitive to the NTD-related nutrients, that also had a previous link to NTDs). Our rationale to limit our focus to miRNAs with a previous NTD link when constructing nutrient-sensitive miRNA regulatory networks was driven by the high number of miRNAs associated with the nutrients of interest. For example, approximately 1000 miRNAs were identified to be sensitive to each of choline, inositol and iron. Lastly, the targetome files were used to construct nutrient-sensitive miRNA-DEG regulatory networks in Cytoscape (version 3.8.2)^32^.

### Nutrient-sensitive transcription factor regulatory network analysis

Next, we used iRegulon (version 1.3) to identify possible transcriptional regulators (TRs) predicted to co-regulate DEGs in cases^40^. The iRegulon software uses a reverse engineering approach to predict the transcriptional regulatory network of a given gene list, using *cis*-regulatory sequence analysis, and has been shown to outperform other motif-discovery methods^40^. First, a whole genome ranking of 22,284 human genes (RefSeq) for each regulatory motif is generated. Specifically, TR binding motifs, or CHIP-seq peaks (derived from ChIP-seq data sets, also referred to as tracks), were identified in the regulatory sequences in the 20 kb (kilobase; 1000 base pairs of DNA) surrounding the transcription start site (TSS) of each gene (from −10 kb to +10 kb of the TSS)^40^. Then, the list of differentially expressed genes is input and tested against the ranked gene list using an area under the cumulative recovery curve (AUC). AUC scores are normalized to normalized enrichment scores (NES). Motifs with high NES scores represent motifs that have a large proportion of the differentially expressed genes (input genes) within its top rankings. We used a NES threshold of 3.0 to determine motif enrichment, which corresponds to an FDR of 3-9%. At the same time, the subset of genes from the input list that are predicted to be targeted by a given motif is identified using the leading edge of the recovery curve. Lastly, iRegulon uses a *Motif2TR* procedure to link candidate motifs (those with a NES>3.0) with TRs using a constructed database that links 1,191 human TRs to 6,031 motifs (based on motif-TR direct annotations, orthology, and motif-motif similarity)^40^.

Two lists of candidate TRs associated with enriched motifs were produced: One representing TRs targeting up-regulated genes in cases, and the other targeting down-regulated genes in cases. Within these lists, TRs with nutrient cofactors were identified using the same dataset of protein (gene)-nutrient interactions used above, to identify DEGs with nutrient cofactors^31^. Nutrient-sensitive TRs and their direct DEG targets were then exported as network files, separately for up-and down-regulated DEG lists. Nutrient-sensitive TR-DEG regulatory networks were visualised in Cytoscape (version 3.8.2)^32^.

## Results

### Data pre-processing

Expression value distribution, density, and intensity were similar across all nine samples (Supplementary Figure S1A). One case sample (GSM94601) identified as an outlier in UMAP analysis was excluded (Supplementary Figure S1A), consistent with approaches taken in previous analyses of this dataset^13^. The remaining eight samples were then re-clustered using UMAP, and complete separation of the case and control groups was evident (Supplementary Figure S1B). Three cases and five control samples were retained for subsequent analyses.

### Amniocyte gene expression differs in cases compared to controls

In a comparison of global amniocyte gene expression, 1,155 genes (2.11% of probesets sequenced) were differentially expressed between cases and controls (*q*<0.05; absolute log fold change ≥2). After the removal of duplicate probesets and genes with no available Entrez ID (Supplementary Figure S2 and Table S2), 880 DEG (downregulated: n=725, upregulated: n=155) were carried forward for further analyses (Figure 1).

**Figure 1.**
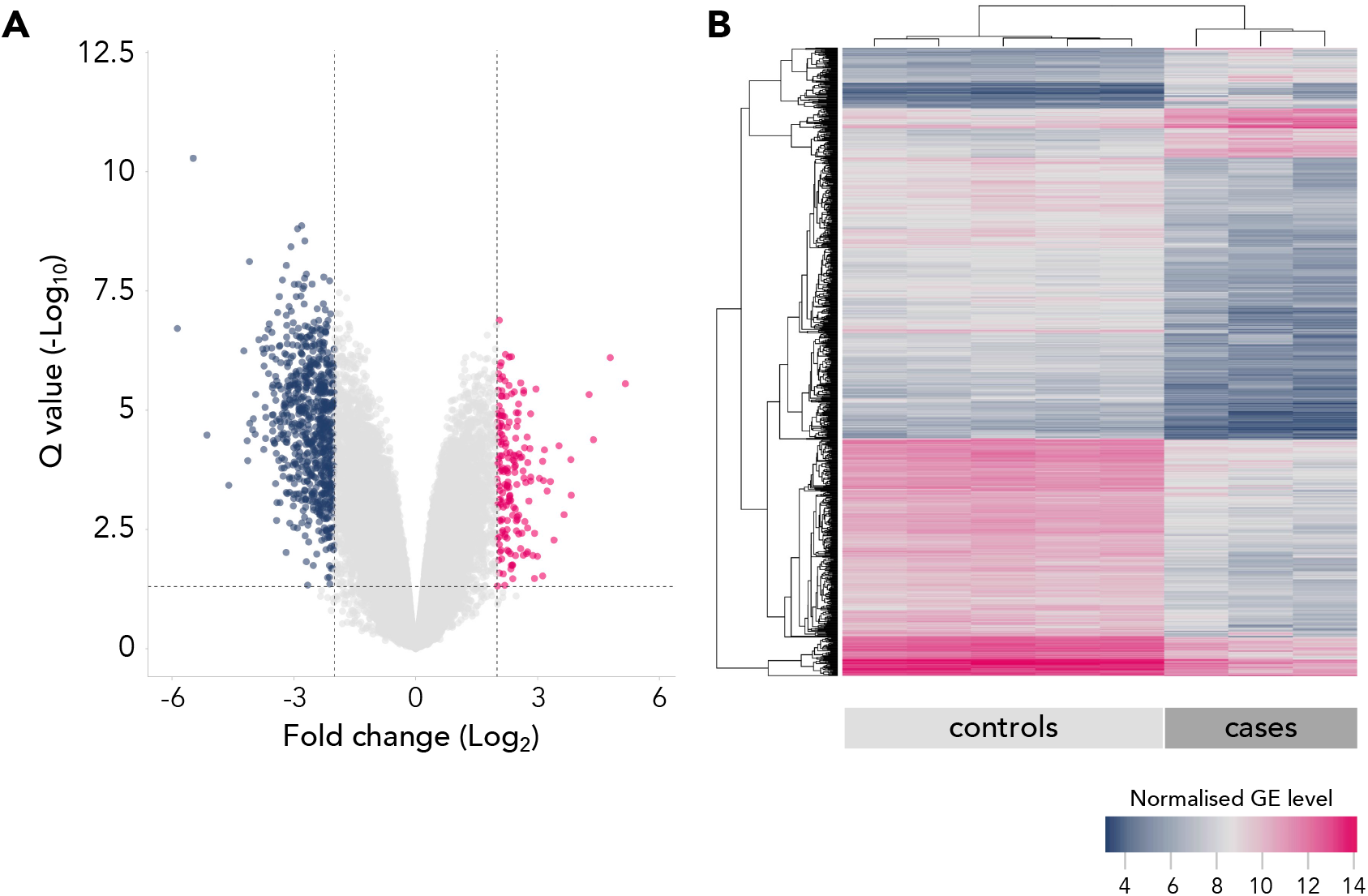
Amniocyte gene expression is different in fetuses with NTDs compared to controls. **(A)** Volcano plot and **(B)** heat map of genes with increased (n=155) and decreased (n=725) expression in cases compared to controls (clustering method: average linkage; distant measurement method: Euclidean; clustering applies to rows and columns). Differential expression was determined at absolute fold change > 2.0 and q < .05. GE = gene expression.

After functional annotation of the DEGs using PANTHER (Supplementary Figure S3), the top identified GO molecular functions of DEGs in cases were binding (n=274 genes), catalytic activity (n=152) and regulatory functions (n=120). The top identified biological processes of DEGs in cases were cellular processes (n=431 genes), biological regulation (n=290), and metabolic processes (n=226). The top protein classes of DEGs were protein modifying enzymes (n=71 genes), gene-specific transcriptional regulators (n=63), and metabolite interconversion enzymes (n=49). The full DEG list is provided in Supplementary Table S3.

### Nutrient-gene relationships identified in genes dysregulated in cases

We next investigated whether DEGs in case amniocytes coded for proteins that had nutrient cofactors. Of the DEGs in cases, 87 (9.89%) had at least one nutrient cofactor (downregulated: n=69, upregulated: n=18; Table 2). Cofactors included calcium (for n=14 genes), chloride (n=1), iron (n=6), heme (n=4), magnesium (n=22), manganese (n=4), S-Adenosyl methionine (n=3), vitamins B2 (n=4), B3 (n=7), B5 (n=2), B6 (n=1), and B9 (folate; n=1), vitamin C (n=1), zinc (n=19), and metal cations (n=9; Figure 2A). In nearly all DEG clusters interacting with a nutrient cofactor, the majority of DEGs were downregulated, apart from DEGs interacting with calcium (9 of 14 were upregulated) and folate (Folate receptor beta [*FOLR2*] was upregulated and the only folate dependent DEG; Figure 2A).

**Figure 2.**
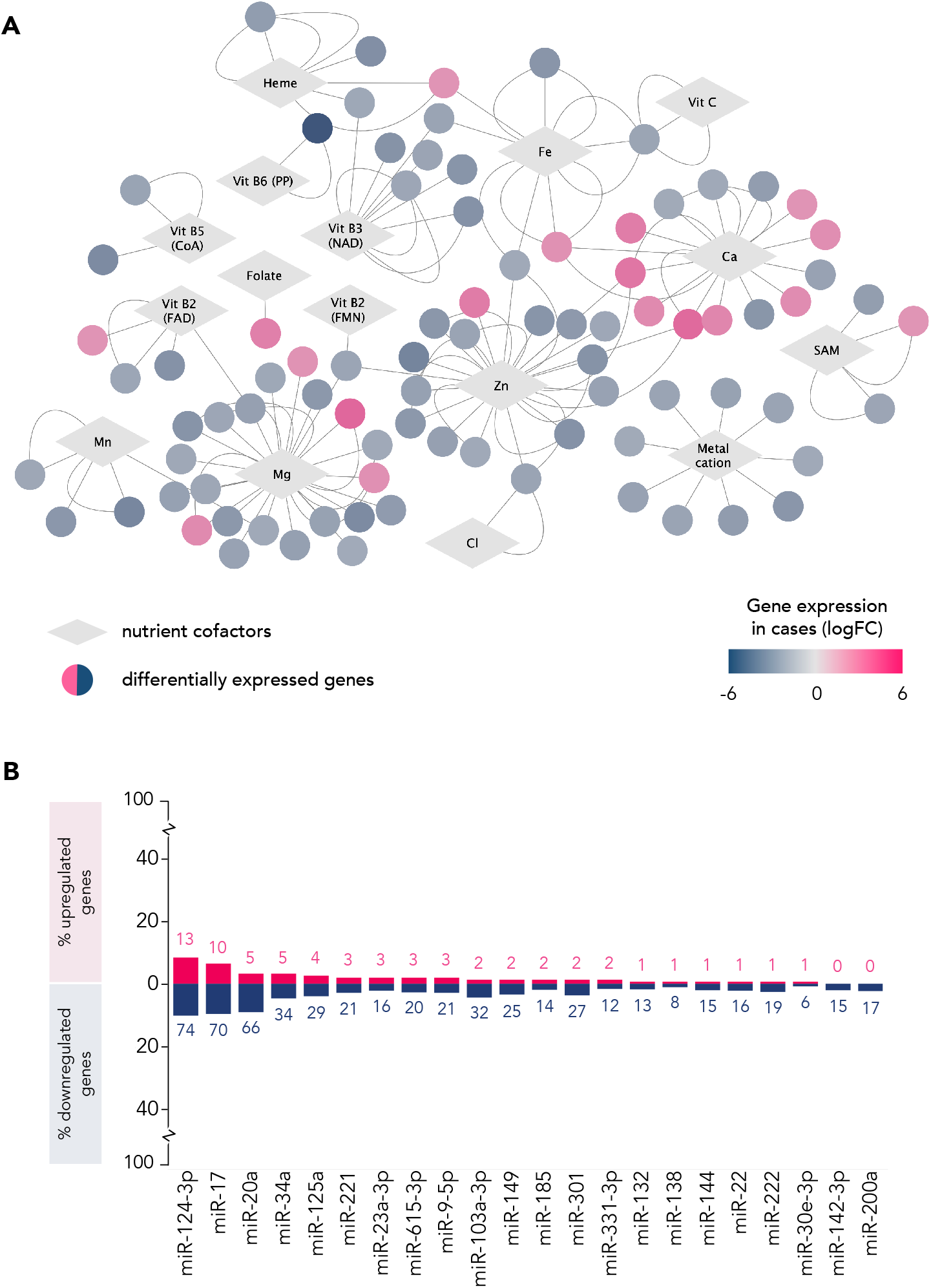
Genes with differential expression in cases compared to controls have (A) nutrient cofactors and (B) are targeted by nutrient-sensitive miRNAs. **(A)** Genes (circle nodes) that had increased (pink) or decreased (blue) expression in cases compared to controls and have a known nutrient cofactor (grey diamonds) are represented in this network diagram. Data on nutrient-gene (protein) interactions was obtained from Scott-Boyer et al. 2016. (B) The largest proportion of DEG are targeted by miR-124-3p, miR-17 (both sensitive to choline, inositol, iron and vitamin D), and miR-20a (sensitive to choline, inositol and iron; miRWalk2.0). Red = upregulated, blue = downregulated, where number = number of DEG targeted by that miRNA. CoA = Coenzyme A. FAD = Flavin adenine dinucleotide. FMN = Flavin mononucleotide. logFC = log fold change. NAD = Nicotinamide adenine dinucleotide. PP = Pyridoxal phosphate. SAM = S-Adenosyl methionine. Vit = vitamin.

Functional annotation of nutrient-dependent DEGs in case amniocytes revealed the top GO molecular functions to be catalytic activity (n=46 genes), binding (n=20), and transporter and molecular transducer activity (n=4 each; Supplementary Figure S4). The top identified biological processes of DEGs with a nutrient cofactor were cellular process (n=51 genes), metabolic process (n=33), and biological regulation (n=33), and the top protein classes were metabolite interconversion enzymes (n=23 genes), protein modifying enzymes (n=14), and calcium-binding proteins (n=6).

The top molecular functions of dysregulated genes with a nutrient cofactor were lipid and purine nucleotide metabolic processes, post-translational protein modifications, and cellular homeostasis and motility-related processes (Table 1). Specifically, the three most downregulated, nutrient-dependent genes in cases were Ethanolamine-Phosphate Phospho-Lyase (*ETNPPL*; vitamin B6-dependent; FC=-5.14, *q*=0.002), involved in lipid metabolism, Baculoviral IAP Repeat Containing 3 (*BIRC3*; zinc-dependent; FC=-3.40, *q*=0.002), associated with apoptosis inhibition, inflammatory signalling pathways, and cell proliferation, and Polypeptide N-Acetylgalactosaminyltransferase 3 (*GALNT3*; manganese-dependent; FC=-3.31, *q*=4.57e^-4^), involved in O-linked oligosaccharide biosynthesis. The three most upregulated genes were Adenylosuccinate Synthase 1 (*ADSSL1*; magnesium-dependent; FC=3.23, *q*=0.009), which codes for an enzyme involved in purine nucleotide synthesis and metabolism, Matrix metallopeptidase 16 (*MMP16*; zinc- and calcium-dependent; FC=3.17, *q*=0.002), involved in extracellular matrix degradation in normal physiological and disease processes, and Ectonucleotide Pyrophosphatase/Phosphodiesterase 2 (*ENPP2*; zinc- and calcium-dependent; FC=2.83, *q*=0.006), associated with motility-related processes, including angiogenesis and neurite outgrowth.

**Table 1.**
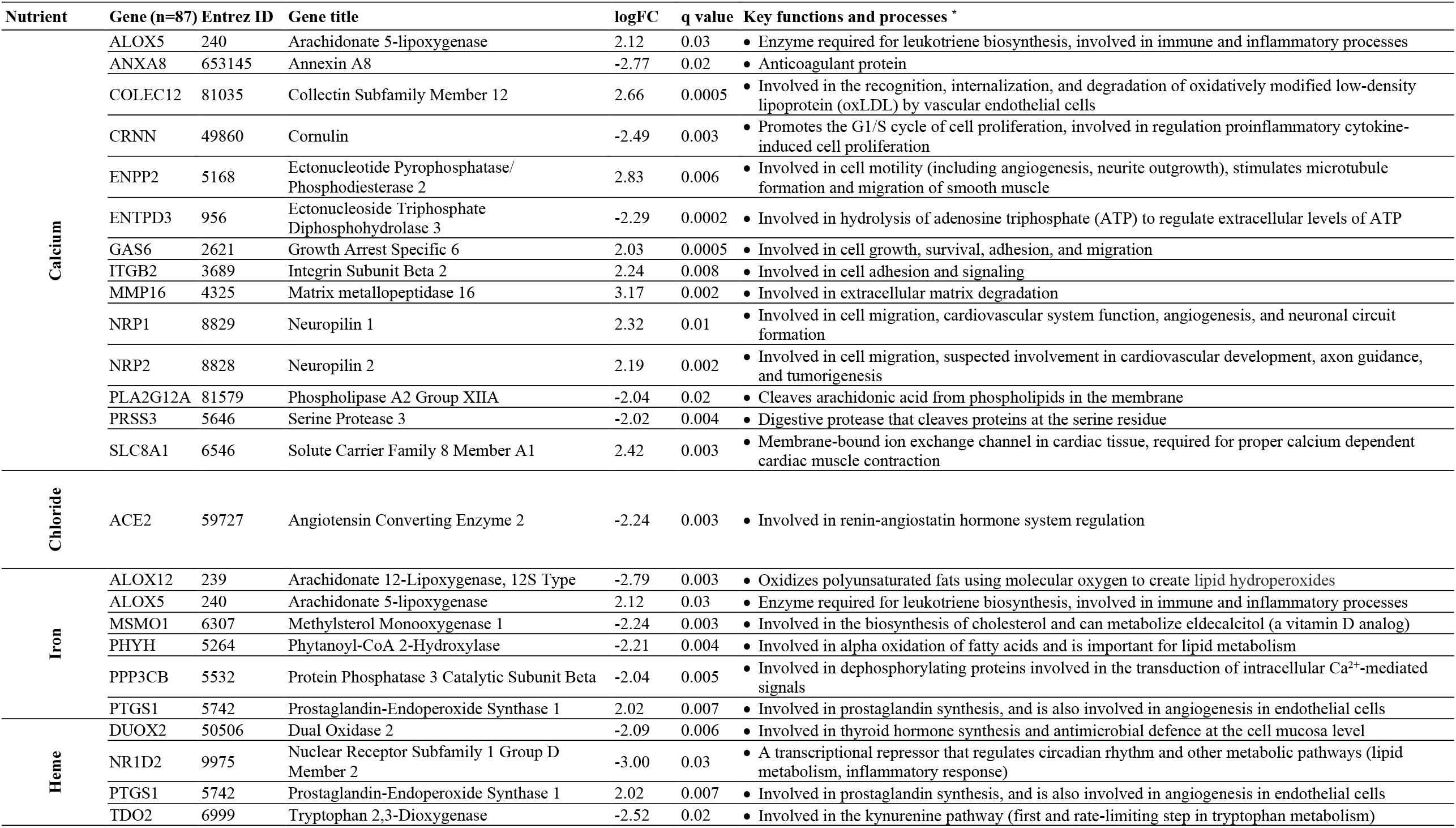
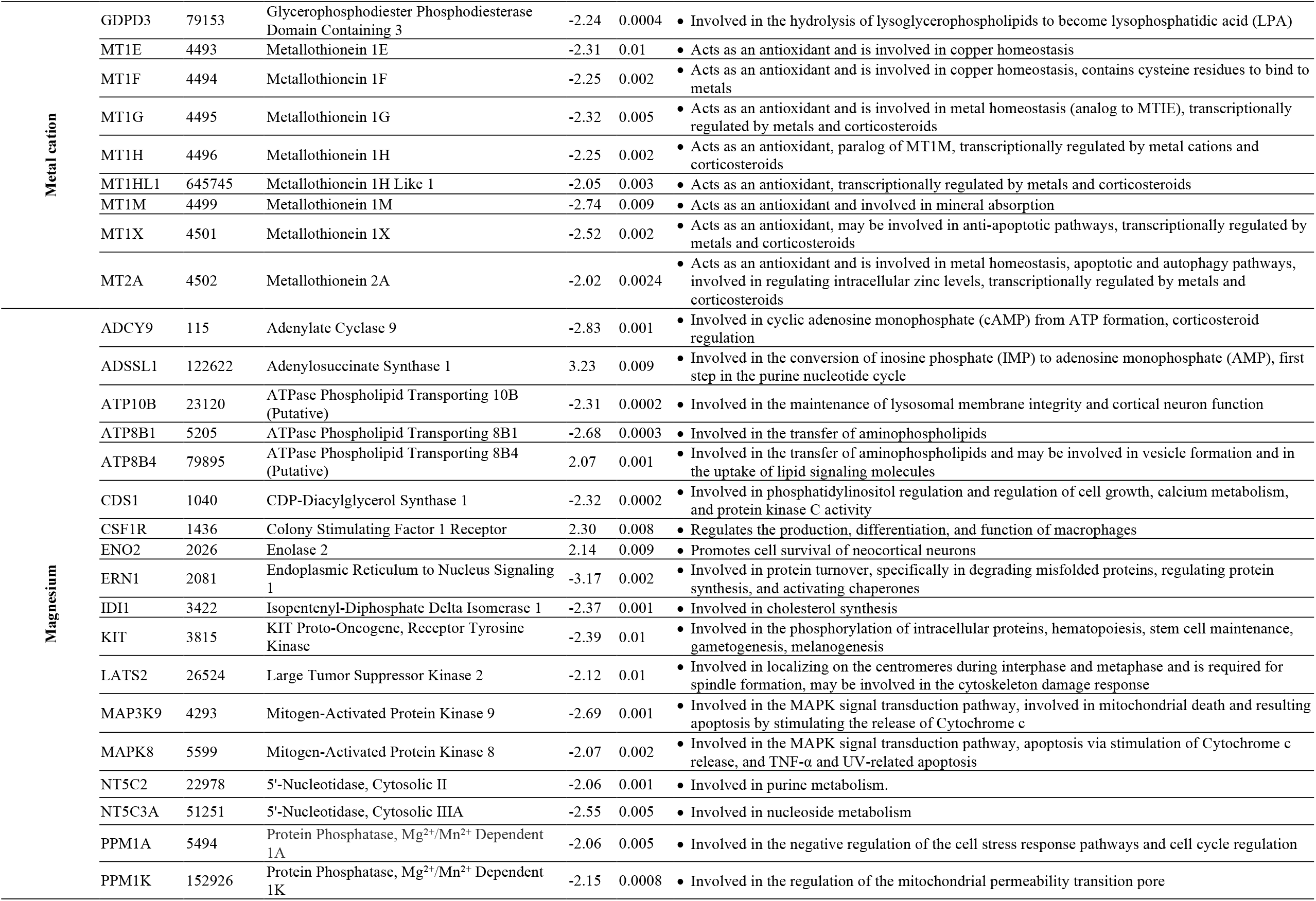
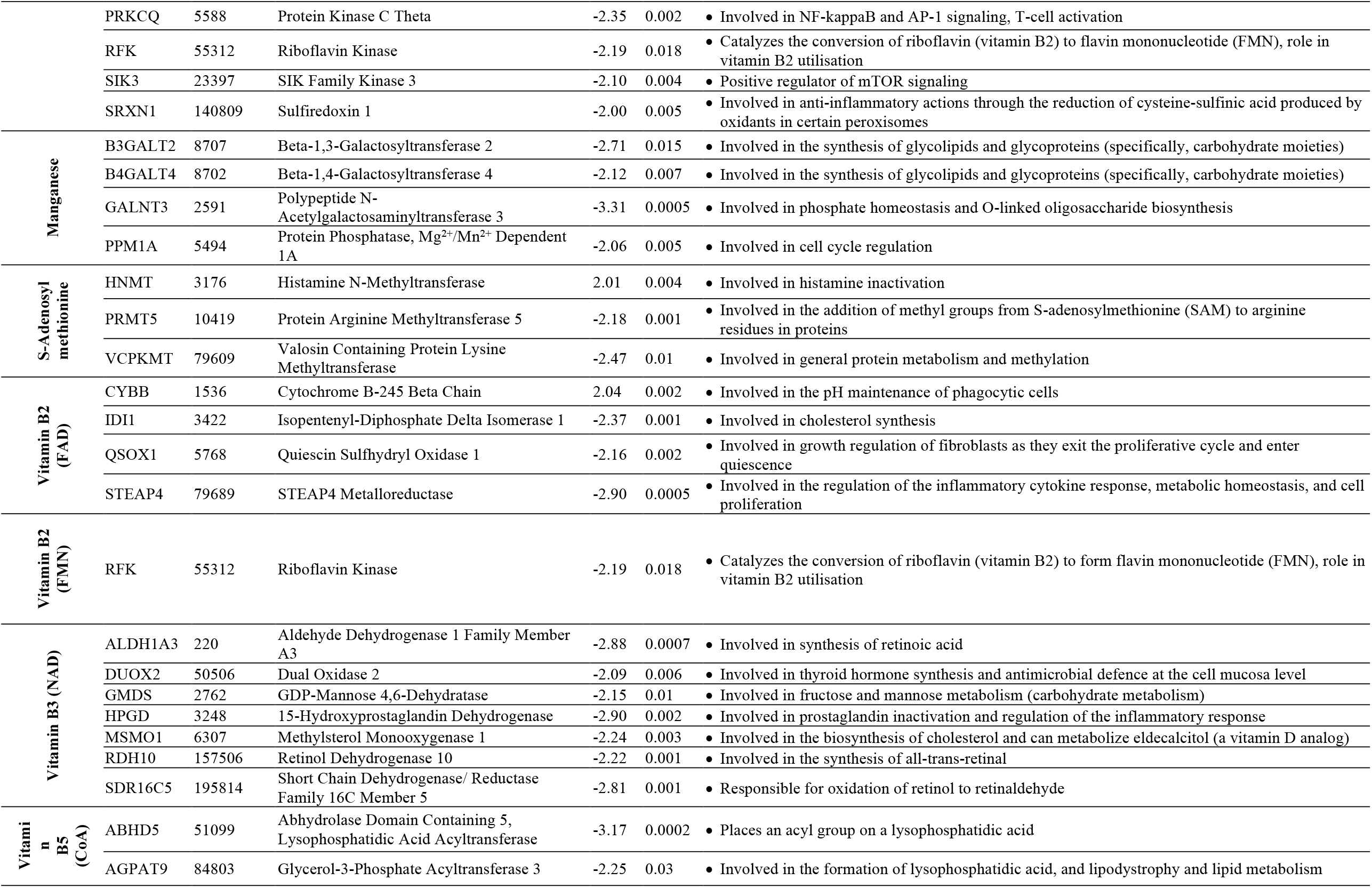
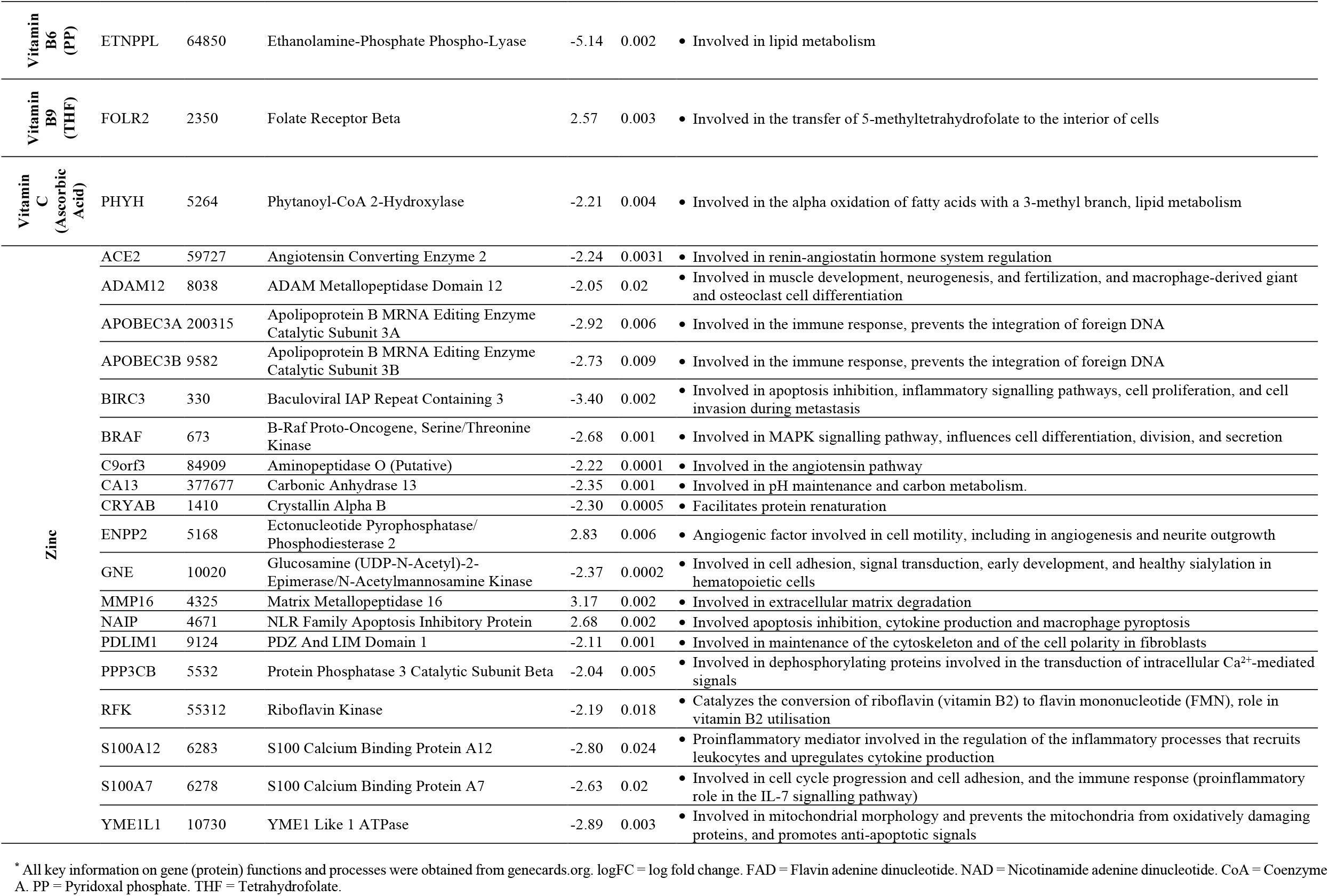
Genes with a nutrient cofactor that were differentially expressed in cases (vs. controls).

Overall, nutrient-dependent, downregulated genes in cases were involved in immune-related processes and nutrient metabolism and homeostasis (Table 1). This included eight genes from the metal cation-binding Metallothionein (*MT*) gene family (*MT1E, MT1F, MT1G, MT1H, MT1HL1, MT1M, MT1X* and *MT2A*), associated with intracellular metal homeostasis, as well as 16 zinc-interacting genes involved in immune and inflammatory processes, antioxidant activity, mitochondrial morphology, the renin-angiotensin pathway, and cell signaling (Table 2). Downregulated genes coding for proteins with B vitamin cofactors included Riboflavin Kinase (*RFK*), which is essential for vitamin B2 utilization, *ETNPPL* and Glycerol-3-Phosphate Acyltransferase 3 (*AGPAT9*), which are involved in lipid metabolism, and Methylsterol Monooxygenase 1 (*MSMO1*) and Isopentenyl-Diphosphate Delta Isomerase 1 (*IDI1*), which are involved in the biosynthesis of cholesterol.

**Table 2.** Transcription factors with a nutrient cofactor that are predicted to target genes that are differentially expressed in cases compared to controls.

| Transcription factor | NES * | # Of targets (DEG) | # Of motifs/ tracks | Cluster code ** | Nutrient cofactor(s) | Linked to NTDs? |
| --- | --- | --- | --- | --- | --- | --- |
| Putatively targeting upregulated genes in cases: |  |  |  |  |  |  |
| *CHURC1* | 3.27 | 18 | 1/0 | M5 | Zinc | No |
| *CTBP2* | 3.13 | 5 | 0/1 | T6 | Vit B3 (NAD) | Yes |
| *EP300* | 3.33 | 9 | 1/0 | M5 | Vit B5 (CoA), Zinc | Yes |
| *MYLK* | 3.28 | 7 | 1/0 | M14 | Mg, Ca | Yes |
| *QSOX1* | 3.69 | 25 | 2/0 | M5 | Vit B2 (FAD) | Yes |
| *RARB* | 3.69 | 17 | 1/0 | M5 | Vit A | Yes |
| *SIRT6* | 3.33 | 9 | 1/0 | M5 | Vit B3 (NAD), Zinc | Yes † |
| *SMAD1* | 3.69 | 25 | 2/0 | M5 | Zinc | Yes |
| *TROVE2* | 3.42 | 35 | 1/0 | M12 | Metal cation | Yes |
| *VDR* | 3.69 | 17 | 1/0 | M5 | Vit D | Yes |
| *YY1* | 3.69 | 17 | 1/0 | M5 | Zinc | Yes |
| Putatively targeting downregulated genes in cases: |  |  |  |  |  |  |
| *EP300* | 3.64 | 302 | 0/3 | T2 | Vit B5 (CoA), Zinc | Yes |
| *SMAD3* | 3.49 | 252 | 5/0 | M1 | Zinc | Yes |
| *SMAD4* | 3.19 | 40 | 1/0 | M8 | Zinc | Yes |
| *TP53* | 3.49 | 252 | 5/0 | M1 | Zinc | Yes |
| *TP63* | 3.49 | 252 | 5/0 | M1 | Zinc | No |
| *TP73* | 3.49 | 252 | 4/0 | M1 | Zinc | Yes |

\* Maximal enrichment score for the motif(s) or track(s) predicted to be targeted by the TF. \*\* Cluster codes indicate the motif (M) or track (T) clusters a TF is associated with. † Previous link to NTDs in animal models only. NES = Normalized enrichment score. DEG = differentially expressed genes.

Upregulated genes in cases included those involved in immune and inflammatory processes, such as Arachidonate 5-lipoxygenase (*ALOX5*; calcium- and iron-dependent), Colony Stimulating Factor 1 Receptor (*CSF1R*; magnesium-dependent), *FOLR2* (folate-dependent), and NLR Family Apoptosis Inhibitory Protein (*NAIP*; zinc-dependent; Table 2). Calcium-dependent Neuropilin 1 (*NRP1*) and Ectonucleotide Pyrophosphatase/Phosphodiesterase 2 (*ENPP2*), and iron- and heme-dependent Prostaglandin-Endoperoxide Synthase 1 *(PTGS1*), were also upregulated in cases and are involved in angiogenesis (Table 1).

### Nutrient-sensitive miRNAs target differentially regulated genes in cases

We identified 22 unique miRNAs that had a previous link to NTDs and were sensitive to the nutrients of interest identified a priori (Figure 2B). Collectively, these 22 miRNAs targeted 39% (n=344) of DEGs in cases (19% [n=155] for B vitamin-sensitive miRNAs, 30% [n=263] for choline-sensitive miRNAs, 23% [n=203] for vitamin D-sensitive miRNAs, 33% [n=289] for inositol sensitive miRNAs, and 33% [n=293] for iron-sensitive miRNAs).

The largest proportion of DEGs were targeted by *MIR-124-3p* (n=87 DEGs), a known suppressor of tumor proliferation and invasion, and neural inflammation^41-43^, with activity sensitive to choline, inositol, iron, and vitamin D. *MIR-17* and *MIR-20a*, both belonging to the miR-17/92 cluster with known roles in cell cycle, proliferation and apoptosis, immune function, and neurodegeneration^44^, targeted the next largest proportions of DEGs in amniocytes of cases (n=80 DEGs for *MIR-17*, sensitive to choline, inositol, iron, and vitamin D; n=71 DEGs for *MIR-20a*; sensitive to choline, inositol and iron; Figure 2B). Two of the miRNAs (*MIR142-3p* and *MIR-200a*; both involved in epithelial-to-mesenchymal transition^45,46^), only targeted downregulated genes in cases, while the remaining 20 miRNAs identified targeted both up- and downregulated DEGs in cases (Figure 2B).

Network analysis revealed that seven of these 22 unique miRNAs were known to be sensitive to only one nutrient of interest. This included inositol-sensitive miRNAs: *MIR-23a-3p, MIR-142-3p, MIR-144, MIR-185*; iron-sensitive miRNAs: *MIR-200a, MIR-301*; and vitamin B6-sensitive miRNA: *MIR-30e-3p* (Figure 3A-B). Iron, inositol, and choline-sensitive miRNAs targeted the largest proportion of DEGs in cases (Figure 3C). miRNA-targetome networks for DEG in amniocytes from cases compared to controls revealed a high degree of overlap in DEGs targeted by the 22 unique nutrient-sensitive miRNAs (Figure 3D).

**Figure 3.**
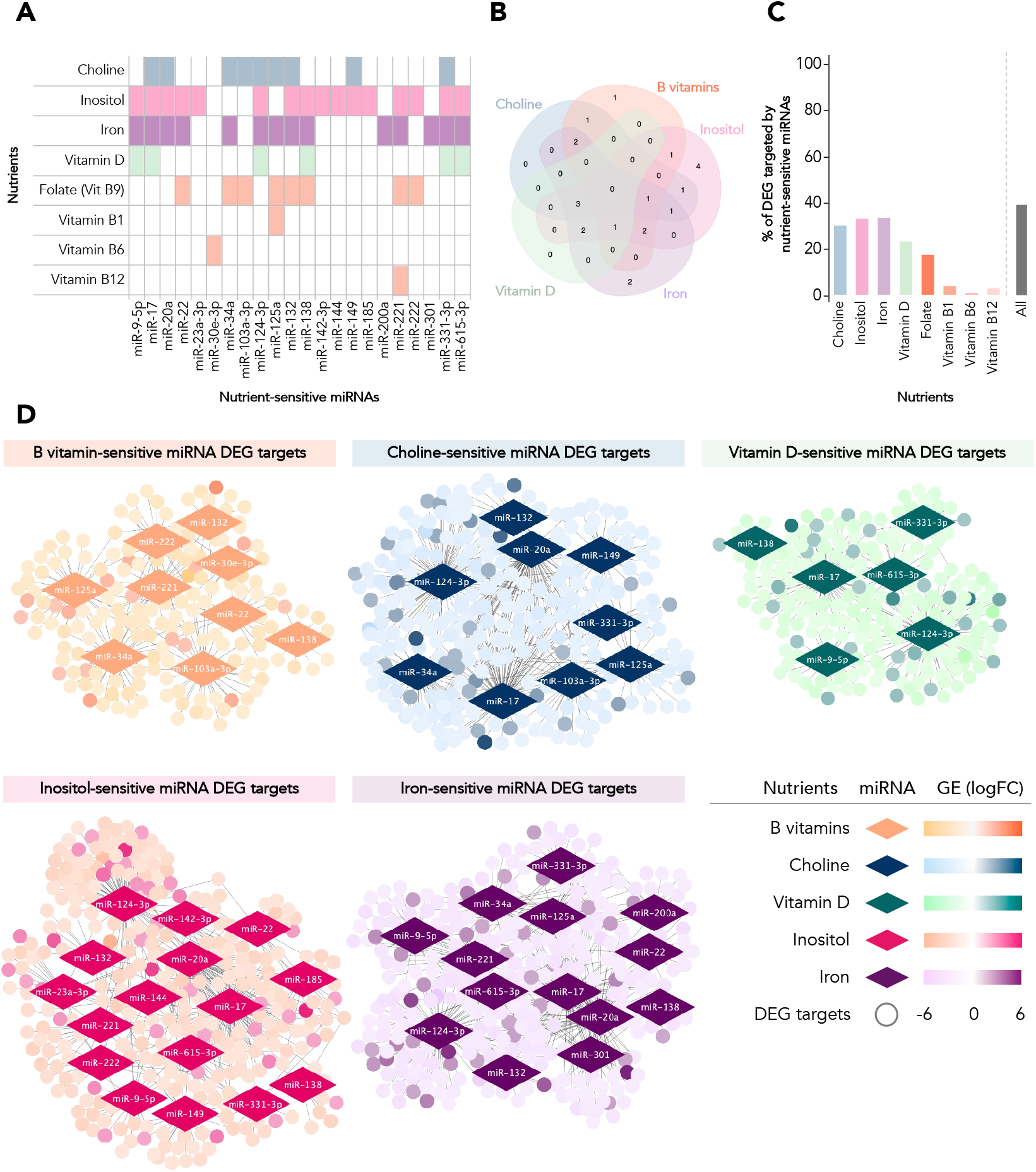
Nutrient-sensitive miRNA-DEG network analysis. (A-B) 22 unique miRNAs known to be sensitive to the activity of nutrients of interest (identified a priori, miRWalk2.0) with a previous link to NTDs. Of the 22 miRNAs, seven were sensitive to only one nutrient of interest (inositol-sensitive: miR-23a-3p, miR-142-3p, miR-144, miR-185; iron-sensitive: miR-200a, miR-301; vitamin B6-sensitive: miR-30e-3p). (C) Iron, inositol and choline-sensitive miRNAs targeted the largest proportion of DEGs in cases, respectively. Collectively, the 22 unique miRNAs targeted 39% of DEG in cases (n=344; B vitamin-sensitive miRNAs: 19% [n=155], choline-sensitive miRNAs: 30% [n=263], vitamin D-sensitive miRNAs: 23% [n=203], inositol-sensitive miRNAs: 33% [n=289], iron-sensitive miRNAs: 33% [n=293]). (D) Nutrient-sensitive miRNA-targetome networks for DEG in cases compared to controls. Diamonds = nutrient-sensitive miRNAs. Circles (nodes) = DEG targets, where lighter shades = downregulated in cases, darker shades = upregulated in cases. DEG = differentially expressed genes. GE = gene expression. FC = fold change. Vit = vitamin.

### Nutrient-dependent transcriptional regulators target differentially expressed genes

iRegulon analysis revealed 125 and 106 unique TRs predicted to target up- and down-regulated genes, respectively, in case amniocytes (Supplementary Table S4). Of these, 16 TRs had nutrient cofactors and are predicted to target down- (n=5) and up-regulated (n=10) DEGs, or both (n=1; Table 2). Nutrient cofactors of these TRs included calcium (for n=1 TR), magnesium (n=1), metal cations (n=1), vitamins A (n=1), B2 (n=1), B3 (n=2), B5 (n=1), D (n=1) and zinc (n=10; Figure 4A). Collectively, these 16 TRs are predicted to regulate the expression of 52% (n=460) of DEGs (Figure 4B). E1A Binding Protein P300 (*EP300*), a TR with known roles in cell growth, differentiation, and NTDs, was predicted to regulate the highest number of DEGs of the TRs identified. *EP300* had 311 expected targets (upregulated: n=9, downregulated: n=302; 35.3% of total DEGs; Table 2), and zinc and vitamin B5 cofactors. The zinc-requiring P53 family proteins (*TP53, TP63*, and *TP73*), involved in cell proliferation and apoptosis, were the next-largest predicted regulators of DEGs, potentially targeting 252 downregulated genes in cases (28.6% of total DEGs).

**Figure 4.**
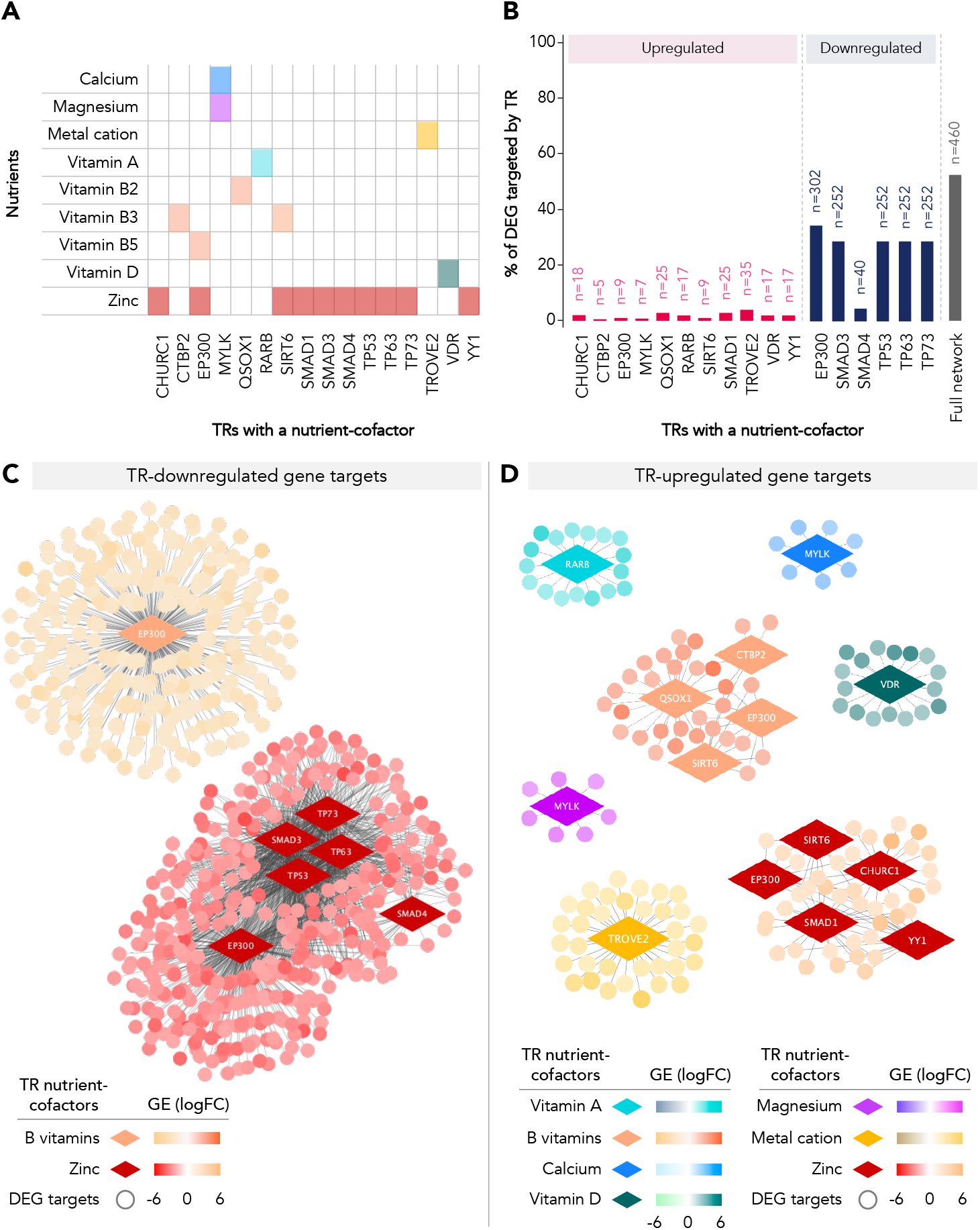
Nutrient-sensitive transcriptional regulatory networks for DEGs in cases compared to controls. **(A)** 16 transcriptional regulators with nutrient-cofactors that are predicted to regulate DNA motifs or tracks of DEG were identified. Nutrient cofactors of these TRs included calcium (for n=1 TR), magnesium (n=1), metal cations (n=1), vitamins A (n=1), B2 (n=1), B3 (n=2), B5 (n=1), D (n=1) and zinc (n=10). **(B)** Collectively, these 16 TRs are predicted to regulate the expression of 52% (n=460) of DEGs. Red = upregulated in cases, blue = downregulated in cases, grey = total proportion of DEG targeted by a nutrient-sensitive TR. Nutrient-sensitive TR-gene networks were visualised for TRs predicted to target **(C)** downregulated genes (n=5 unique TRs, left-bottom panel) and **(D)** upregulated genes (n=10 unique TRs, right-bottom panel) separately. One TR (E1A Binding Protein P300; EP300) is predicated to target both up- and down-regulated genes in cases. Diamonds = nutrient-sensitive TRs. Circles (nodes) = DEG targets, where colours on the left of the colour palette = downregulated in cases, right of the colour palette = upregulated in cases. DEG = differentially expressed genes. GE = gene expression. FC = fold change. TR = transcriptional regulator.

Of TRs identified as potential regulators of upregulated genes, the metal-cation reliant TROVE domain family, member 2 (*TROVE2*) gene, a known regulator of immune and inflammatory processes, was associated with the largest proportion (n=35 [4.0%] upregulated genes in cases). Quiescin Sulfhydryl Oxidase 1 (*QSOX1;* vitamin B2-dependent) which targeted the next-largest group of DEGs (n=25 upregulated genes), was also downregulated in cases (FC=-2.16, *q*=0.002; Table 2). One of the P53 family proteins (*TP63*), as well as an additional zinc-dependent TR identified (Churchill Domain Containing 1 [*CHURC1*]), have not been previously linked to NTDs.

Nutrient-sensitive TR-gene networks were visualised for enriched TRs predicted to target downregulated and upregulated genes (Figure 4C-D). All TRs predicted to target downregulated genes had zinc as a cofactor, and one (*EP300*) was also vitamin B5-dependent (Figure 4C). TRs targeting upregulated genes were dependent on calcium, magnesium, metal cations, vitamins A, B3, B5, D, and zinc (Figure 4D). In the TR-gene network figures, it is visually evident that the regulons of downregulated genes targeted by nutrient-sensitive TRs were much larger than the upregulated regulons, but were fewer in number (Figure 4C-D).

## Discussion

Using a nutrient-focused gene expression analysis pipeline, we identified multiple nutrient-sensitive genes and gene regulatory networks that were dysregulated in amniocytes in a small cohort of fetuses with NTDs. We found that over half of the dysregulated genes were predicted to be co-regulated by TRs and miRNAs that are sensitive to, or dependent on, nutrients with known roles in NTD etiology, namely zinc, B vitamins, and B vitamin-like nutrients (choline and inositol). These findings generate new hypotheses on the role of nutrient-gene interactions in NTDs, and suggest that nutrient-sensitive genes and gene regulatory networks may explain an important portion of gene expression signatures associated with NTDs. Profiling nutrient-transcriptome interaction networks in the context of NTDs may help to illustrate, in part, the gene-environment interactions that characterise an intrauterine environment that is permissive of NTDs and associated comorbidities.

Several zinc-dependent genes and gene regulatory networks were dysregulated in case amniocytes compared to controls. Zinc is a key regulator of gene expression, and is essential for the rapid DNA synthesis, cell growth, and proliferation that accompany embryonic development, including neural tube closure^47,48^. Low maternal zinc status may be associated with increased NTD risk^9^, and zinc imbalance in animal studies leads to NTDs, possibly through modulation of *p53* activity^49^. Here, three zinc-dependent P53 family proteins were identified as transcriptional regulators predicted to target over a quarter of DEGs in case amniocytes, and eight Metallothionein genes, which are essential for maintaining zinc homeostasis and controlling zinc-dependent cellular signaling^50^, were downregulated in cases. Altogether, our findings support a role for zinc-associated gene dysregulation in NTD genetic phenotypes, in alignment with previous animal studies, and illustrate novel zinc-dependent gene networks in an NTD-affected a human cohort.

One zinc- and vitamin B5-dependent TR, *EP300*, was predicted to target over one third of DEGs in case amniocytes. *EP300* has been studied in mouse models of NTDs and is predicted to activate pathways that promote fusion of the neural tube^51,52^. In humans, rare deleterious variants in the *EP300* gene have recently been associated with NTDs^53^, and here, *EP300* was the dominant, nutrient-sensitive focal point for DEG regulation in cases. Two additional nutrient-dependent TRs identified here further support a role for both zinc and B vitamin-interacting gene dysregulation in cases: Sirtuin 6 (*SIRT6*; vitamin B3- and zinc-dependent) and *CHURC1* (zinc-dependent). *SIRT6* suppression by high glucose levels is a proposed to mechanism linking maternal diabetes to increased risk of fetal NTDs^54^, however, *CHURC1* has not previously been associated with NTDs. In zebrafish and xenopus, *churc1* is known to play a role in embryogenesis and neuroectodermal development^55,56^. In humans, *CHURC1* has been investigated in relation to autism, where a microdeletion in a gene region containing the genes for both *CHURC1* and methylenetetrahydrofolate dehydrogenase 1 (*MTHFD1*), a folate metabolism enzyme with known links to NTD etiology^11,57^, associated with increased autism risk^58^. While research on zinc-B vitamins interactions is limited, there is some evidence that zinc may have a role in niacin (vitamin B3) metabolism through interaction with pyridoxine (vitamin B6)^59,60^. Overall, the presence of dysregulated zinc- and B vitamin-dependent gene networks in our results suggest that both zinc- and B vitamin-gene interactions in NTDs should be targets of further study.

To date, folates (vitamin B9), and more recently, folate one-carbon metabolism (OCM) cofactors (namely vitamins B12 and B6), have been the central nutrient targets for NTD prevention. In agreement with the established role of folates and OCM cofactors in NTDs, we identified nine miRNAs with known associations to NTDs that were also B vitamin-sensitive, eight being sensitive to folate, and were predicted to regulate the expression of nearly one fifth of DEGs in case amniocytes. Further, the three miRNAs that targeted the largest proportion of DEGs in cases (*MIR-124-3p, MIR-17*, and *MIR-20a*) were each sensitive to both choline and inositol, water soluble B vitamin-like molecules with known roles in NTD etiology^6,8^, and four TRs with B vitamin cofactors (*EP300* and *SIRT6*, as well as C-terminal-binding protein 2 [*CTBP2*] and *QSOX1*) predicted to target DEGs in cases were identified. Both *QSOX1* and *CTBP2* have inferred associations with NTDs via their interactions with valproic acid and folic acid^61^, and the role of the *CTBP2* gene in the Wnt and Notch signaling pathways^62^. However, no studies have provided direct evidence linking *QSOX1* or *CTBP2* to NTD pathogenesis or phenotype in humans, and here, *QSOX1* expression was also downregulated in case amniocytes compared to controls. Taken together, our findings advance beyond a folic acid (vitamin B9)-centric view of the role of B vitamins in NTDs and highlight B vitamin-sensitive or dependent gene candidates for further study in NTDs.

Additionally, we found that six miRNAs with known associations to NTDs were vitamin D-sensitive, and collectively, were predicted to target nearly one quarter of DEGs in case amniocytes. Vitamin D receptor (VDR) was also identified as a transcription factor targeting DEGs, and Methylsterol monooxygenase 1 (*MSMO1*), associated with vitamin D analog metabolism, was downregulated in cases. Vitamin D plays important anti-inflammatory and immunomodulatory roles^63^, and low maternal vitamin D levels have been associated with fetal NTDs^7^. In mice, vitamin D3 supplementation prevents lipopolysaccharide-induced NTDs via increased folate transport from the maternal circulation to the embryo^64^. Thus, our findings support vitamin D as a nutrient of interest for further research on nutrient-gene interactions in NTDs.

Maternal immune dysregulation is also known to associate with an increased risk of fetal NTDs^65,66^, and here, several nutrient-dependent DEGs in cases were involved in inflammatory and immune-related biological processes. While it is possible that the dysregulated amniocyte gene expression in immune or inflammatory process-related genes was shaped by maternal health factors, activation of pro-inflammatory signaling pathways at the site of improper neural tube closure has also been reported in fetuses with spina bifida^67,68^, and may serve as an additional ‘hit’ to the developing central nervous system. Whether signaling changes in spinal cord injury-induced secondary lesion cascades could influence gene expression in amniocytes is an area requiring further study. Given the relationships between nutrient intakes and inflammatory load^69^, and the importance of nutrition in supporting immune signaling and functions^70^, the interactions between nutrient-sensitive and immune/inflammatory processes in NTD pathogenesis and phenotype is an area requiring further investigation.

The small size of this cohort is a key limitation to our study that may limit the validity of our findings. However, the rarity of NTDs and the challenges that accompany amniotic fluid collection make these data a valuable resource for generating new hypotheses on the role of nutrient-gene interactions in NTDs. While amniocentesis was performed after the period of neural tube closure, the gene expression changes observed may still reflect a uterine environment that was permissive of altered embryonic development. Alternatively, gene expression changes in amniocytes may also be influenced by signaling changes following improper neural tube closure, which could inform on NTD phenotype and the uterine environment that is permissive of comorbidities that develop later in pregnancy. Notably, maternal cell contamination in amniotic fluid is known to occur^71^ and could have confounded in the amniocyte gene expression profiles reported here. To address this limitation and characterise the transcriptome of certain minor cell populations in amniotic fluid, future studies on new samples should consider single-cell type RNA sequencing (sctRNA-seq) and computational approaches applied to sctRNA-seq to detect and control for changes in transcript expression due to off-target cell contamination^72^. Data on cohort characteristics were also limited, and it is plausible that there are unexplored variables that could help explain the differences observed here. These variables may include maternal medical history, or clinical or demographic characteristics, as well as dietary intakes; namely of nutrients that have known associations with NTD pathogenesis. Fortification of wheat products with folic acid had also not been introduced at the time of data collection, and while the original authors did report that all study participants were taking a multivitamin supplement throughout pregnancy^14^, it remains difficult to speculate about the nutrient status, including folate, of the study’s participants. Maternal hyperhomocysteinemia and C677T polymorphism in the *MTHFR* gene are also known risk factors for fetal NTDs^12,73^, and may have influenced the gene expression in cases if present, even if mothers were nutritionally replete. However, maternal biomarker and genotype data were not available for this cohort, limiting our ability to assess relationships between maternal genotype or plasma markers and amniocyte gene expression. Our ability to identify genes encoding for a nutrient-interacting protein was limited by the data available on cofactor-protein interactions, which may have led to an over-representation of well-studied nutrients^31^. Further, as the transcriptional regulators identified through iRegulon to target DEGs are predicted, and include both validated and unvalidated regulatory relationships, causal relationships cannot be determined at this time. Our scope for miRNA-targetome analysis was narrowed to focus on only miRNAs with a previous link to NTDs. This may have prevented the identification of novel miRNAs associated with NTDs, and future research with higher computational resources should consider an expanded focus. Ultimately, these findings can inform testable hypotheses and should be next investigated in experimental models, that could evaluate causal relationships between the bioavailability of multiple micronutrients and resulting changes in gene expression. Confirming these findings in an independent cohort would also be an important next step to replicate and validate these findings.

Building on the work of authors who previously analysed these data to understand gene- and gene pathway dysregulation in isolated NTDs^13,14^, our nutrient-focused network analysis approach integrates knowledge on nutrient-gene interactions with gene expression network analysis to examine the functional networks underlying NTD phenotypes. This is a first step in generating testable hypotheses for better understanding relationships between the bioavailability of, and interactions between, multiple micronutrients and gene expression in NTD pathogenesis or comorbidities. Through a network medicine approach, ‘big’ biomedical data can be leveraged to identify the functional networks that drive health and disease states^15^, and here, we apply this approach in the context of nutrient-gene interactions in NTDs. The nutrient-focused gene expression analysis pipeline applied here allows for the characterisation of nutrient-sensitive mechanisms that underly disease processes, which may aid in the identification of novel nutrition targets for the prevention of disease and promotion of optimal health and developmental outcomes.

## Data Availability

The microarray data used in this analysis were obtained from GEO (GSE4182). The code used for microarray pre-processing has been deposited in a GitHub repository (https://github.com/marina-white/ntd_af_nutrient-gene_study.git).

## Acknowledgments

We thank the authors who originally sequenced these data for making them publicly available in GEO.

## References

1. Christianson A, Howson CP, Modell B. Global report on birth defects: The hidden toll of dying and disabled children. 2006. https://www.marchofdimes.org/mission/march-of-dimes-global-report-on-birth-defects.aspx

2. Copp AJ, Stanier P, Greene NDE. Genetic Basis of Neural Tube Defects. Textbook of Pediatric Neurosurgery. Srpinger, Cham.; 2017:1–28:chap Genetic Basis of Neural Tube Defects.

3. White M, Grynspan D, Van Mieghem T, Connor KL. Isolated fetal neural tube defects associate with increased risk of placental pathology: Evidence from the Collaborative Perinatal Project. Placenta. 10 2021;114:56–67. doi:10.1016/j.placenta.2021.08.052

4. Honnebier WJ, Swaab DF. The Influence of Anencephaly Upon Intrauterine Growth of Fetus and Placenta and Upon Gestation Length. BJOG: An International Journal of Obstetrics and Gynaecology. 1973;80(7):577–588. doi:10.1111/j.1471-0528.1973.tb16030.x

5. Finnell RH, Caiaffa CD, Kim SE, et al. Gene Environment Interactions in the Etiology of Neural Tube Defects. Front Genet. 2021;12:659612. doi:10.3389/fgene.2021.659612

6. D’Souza SW, Copp AJ, Greene NDE, Glazier JD. Maternal Inositol Status and Neural Tube Defects: A Role for the Human Yolk Sac in Embryonic Inositol Delivery? Adv Nutr. Feb 1 2021;12(1):212–222. doi:10.1093/advances/nmaa100

7. Nasri K, Ben Fradj MK, Feki M, et al. Maternal 25-hydroxyvitamin D level and the occurrence of neural tube defects in Tunisia. Int J Gynaecol Obstet. Aug 2016;134(2):131–4. doi:10.1016/j.ijgo.2016.01.014

8. Shaw GM, Carmichael SL, Yang W, Selvin S, Schaffer DM. Periconceptional dietary intake of choline and betaine and neural tube defects in offspring. Am J Epidemiol. Jul 15 2004;160(2):102–9. doi:10.1093/aje/kwh187

9. Dey AC, Shahidullah M, Mannan MA, Noor MK, Saha L, Rahman SA. Maternal and neonatal serum zinc level and its relationship with neural tube defects. J Health Popul Nutr. Aug 2010;28(4):343–50. doi:10.3329/jhpn.v28i4.6040

10. De Wals P, Tairou F, Van Allen MI, et al. Reduction in Neural-Tube Defects after Folic Acid Fortification in Canada. New England Journal of Medicine. 2007;357(2):135–142. doi:10.1056/NEJMoa067103

11. Bassuk AG, Kibar Z. Genetic basis of neural tube defects. Semin Pediatr Neurol. Sep 2009;16(3):101–10. doi:10.1016/j.spen.2009.06.001

12. Yan L, Zhao L, Long Y, et al. Association of the maternal MTHFR C677T polymorphism with susceptibility to neural tube defects in offsprings: evidence from 25 case-control studies. PLoS One. 2012;7(10):e41689. doi:10.1371/journal.pone.0041689

13. Li Z, Feng J, Yuan Z. Key Modules and Hub Genes Identified by Coexpression Network Analysis for Revealing Novel Biomarkers for Spina Bifida. Front Genet. 2020;11:583316. doi:10.3389/fgene.2020.583316

14. Nagy GR, Gyõrffy B, Galamb O, Molnár B, Nagy B, Papp Z. Use of routinely collected amniotic fluid for whole-genome expression analysis of polygenic disorders. Clin Chem. Nov 2006;52(11):2013–20. doi:10.1373/clinchem.2006.074971

15. Sonawane AR, Weiss ST, Glass K, Sharma A. Network Medicine in the Age of Biomedical Big Data. Front Genet. 2019;10:294. doi:10.3389/fgene.2019.00294

16. Kang JH, Park HJ, Jung YW, et al. Comparative Transcriptome Analysis of Cell-Free Fetal RNA from Amniotic Fluid and RNA from Amniocytes in Uncomplicated Pregnancies. PLoS One. 2015;10(7):e0132955. doi:10.1371/journal.pone.0132955

17. Zwemer LM, Bianchi DW. The amniotic fluid transcriptome as a guide to understanding fetal disease. Cold Spring Harb Perspect Med. Feb 13 2015;5(4)doi:10.1101/cshperspect.a023101

18. Paoloni-Giacobino A, Grimble R, Pichard C. Genetics and nutrition. Clin Nutr. Oct 2003;22(5):429–35. doi:10.1016/s0261-5614(03)00064-5

19. Bentley TG, Willett WC, Weinstein MC, Kuntz KM. Population-level changes in folate intake by age, gender, and race/ethnicity after folic acid fortification. Am J Public Health. Nov 2006;96(11):2040–7. doi:10.2105/AJPH.2005.067371

20. Ray JG, Wyatt PR, Thompson MD, et al. Vitamin B12 and the risk of neural tube defects in a folic-acid-fortified population. Epidemiology. May 2007;18(3):362–6. doi:10.1097/01.ede.0000257063.77411.e9

21. Belkacemi L, Nelson DM, Desai M, Ross MG. Maternal undernutrition influences placental-fetal development. Biol Reprod. Sep 2010;83(3):325–31. doi:10.1095/biolreprod.110.084517

22. Barrett T, Wilhite SE, Ledoux P, et al. NCBI GEO: archive for functional genomics data sets--update. Nucleic Acids Res. Jan 2013;41(Database issue):D991–5. doi:10.1093/nar/gks1193

23. Gautier L, Cope L, Bolstad BM, Irizarry RA. affy--analysis of Affymetrix GeneChip data at the probe level. Bioinformatics. Feb 12 2004;20(3):307–15. doi:10.1093/bioinformatics/btg405

24. Irizarry RA, Hobbs B, Collin F, et al. Exploration, normalization, and summaries of high density oligonucleotide array probe level data. Biostatistics. Apr 2003;4(2):249–64. doi:10.1093/biostatistics/4.2.249

25. McInnes L, Healy J, Saul N, Lukas G. UMAP: Uniform Manifold Approximation and Projection. Journal of Open Source Software. 2018;3(29):861. doi:https://doi.org/10.21105/joss.00861

26. Stalteri MA, Harrison AP. Interpretation of multiple probe sets mapping to the same gene in Affymetrix GeneChips. BMC Bioinformatics. Jan 15 2007;8:13. doi:10.1186/1471-2105-8-13

27. Maglott D, Ostell J, Pruitt KD, Tatusova T. Entrez Gene: gene-centered information at NCBI. Nucleic Acids Res. Jan 2011;39(Database issue):D52–7. doi:10.1093/nar/gkq1237

28. Ritchie ME, Phipson B, Wu D, et al. limma powers differential expression analyses for RNA-sequencing and microarray studies. Nucleic Acids Res. Apr 2015;43(7):e47. doi:10.1093/nar/gkv007

29. Benjamini Y, Hochberg Y. Controlling the false discovery rate: a practical and powerful approach to multiple testing. Journal of the Royal Statistical Society Series B. 1995;57:289–300.

30. Mi H, Muruganujan A, Casagrande JT, Thomas PD. Large-scale gene function analysis with the PANTHER classification system. Nat Protoc. Aug 2013;8(8):1551–66. doi:10.1038/nprot.2013.092

31. Scott-Boyer MP, Lacroix S, Scotti M, Morine MJ, Kaput J, Priami C. A network analysis of cofactor-protein interactions for analyzing associations between human nutrition and diseases. Sci Rep. Jan 18 2016;6:19633. doi:10.1038/srep19633

32. Shannon P, Markiel A, Ozier O, et al. Cytoscape: a software environment for integrated models of biomolecular interaction networks. Genome Res. Nov 2003;13(11):2498–504. doi:10.1101/gr.1239303

33. Petersen JM, Parker SE, Crider KS, Tinker SC, Mitchell AA, Werler MM. One-Carbon Cofactor Intake and Risk of Neural Tube Defects Among Women Who Meet Folic Acid Recommendations: A Multicenter Case-Control Study. Am J Epidemiol. 06 01 2019;188(6):1136–1143. doi:10.1093/aje/kwz040

34. Kerns JC, Gutierrez JL. Thiamin. Adv Nutr. 03 2017;8(2):395–397. doi:10.3945/an.116.013979

35. Lyon P, Strippoli V, Fang B, Cimmino L. B Vitamins and One-Carbon Metabolism: Implications in Human Health and Disease. Nutrients. Sep 19 2020;12(9)doi:10.3390/nu12092867

36. Chandler AL, Hobbs CA, Mosley BS, et al. Neural tube defects and maternal intake of micronutrients related to one-carbon metabolism or antioxidant activity. Birth Defects Res A Clin Mol Teratol. Nov 2012;94(11):864–74. doi:10.1002/bdra.23068

37. D’Souza SW, Copp AJ, Greene NDE, Glazier JD. Maternal Inositol Status and Neural Tube Defects: A Role for the Human Yolk Sac in Embryonic Inositol Delivery? Adv Nutr. Sep 2020;doi:10.1093/advances/nmaa100

38. Sticht C, De La Torre C, Parveen A, Gretz N. miRWalk: An online resource for prediction of microRNA binding sites. PLoS One. 2018;13(10):e0206239. doi:10.1371/journal.pone.0206239

39. Huang HY, Lin YC, Li J, et al. miRTarBase 2020: updates to the experimentally validated microRNA-target interaction database. Nucleic Acids Res. 01 08 2020;48(D1):D148–D154. doi:10.1093/nar/gkz896

40. Janky R, Verfaillie A, Imrichová H, et al. iRegulon: from a gene list to a gene regulatory network using large motif and track collections. PLoS Comput Biol. Jul 2014;10(7):e1003731. doi:10.1371/journal.pcbi.1003731

41. Huang S, Ge X, Yu J, et al. Increased miR-124-3p in microglial exosomes following traumatic brain injury inhibits neuronal inflammation and contributes to neurite outgrowth. FASEB J. 01 2018;32(1):512–528. doi:10.1096/fj.201700673R

42. Wang Y, Chen L, Wu Z, et al. miR-124-3p functions as a tumor suppressor in breast cancer by targeting CBL. BMC Cancer. 11 15 2016;16(1):826. doi:10.1186/s12885-016-2862-4

43. Majid A, Wang J, Nawaz M, et al. miR-124-3p Suppresses the Invasiveness and Metastasis of Hepatocarcinoma Cells. Front Mol Biosci. 2020;7:223. doi:10.3389/fmolb.2020.00223

44. Mogilyansky E, Rigoutsos I. The miR-17/92 cluster: a comprehensive update on its genomics, genetics, functions and increasingly important and numerous roles in health and disease. Cell Death Differ. Dec 2013;20(12):1603–14. doi:10.1038/cdd.2013.125

45. Eades G, Yao Y, Yang M, Zhang Y, Chumsri S, Zhou Q. miR-200a regulates SIRT1 expression and epithelial to mesenchymal transition (EMT)-like transformation in mammary epithelial cells. J Biol Chem. Jul 22 2011;286(29):25992–6002. doi:10.1074/jbc.M111.229401

46. Xie N, Meng Q, Zhang Y, et al. MicroRNA-142-3p suppresses cell proliferation, invasion and epithelial-to-mesenchymal transition via RAC1-ERK1/2 signaling in colorectal cancer. Mol Med Rep. Aug 2021;24(2)doi:10.3892/mmr.2021.12207

47. Cousins RJ. A role of zinc in the regulation of gene expression. Proc Nutr Soc. May 1998;57(2):307–11. doi:10.1079/pns19980045

48. MacDonald RS. The role of zinc in growth and cell proliferation. J Nutr. 05 2000;130(5S Suppl):1500S–8S. doi:10.1093/jn/130.5.1500S

49. Kakebeen AD, Niswander L. Micronutrient imbalance and common phenotypes in neural tube defects. Genesis. Nov 2021;59(11):e23455. doi:10.1002/dvg.23455

50. Krezel A, Maret W. The Functions of Metamorphic Metallothioneins in Zinc and Copper Metabolism. Int J Mol Sci. Jun 09 2017;18(6)doi:10.3390/ijms18061237

51. Bhattacherjee V, Horn KH, Singh S, Webb CL, Pisano MM, Greene RM. CBP/p300 and associated transcriptional coactivators exhibit distinct expression patterns during murine craniofacial and neural tube development. Int J Dev Biol. 2009;53(7):1097–104. doi:10.1387/ijdb.072489vb

52. Lee S, Gleeson JG. Closing in on Mechanisms of Open Neural Tube Defects. Trends Neurosci. 07 2020;43(7):519–532. doi:10.1016/j.tins.2020.04.009

53. Au KS, Hebert L, Hillman P, et al. Human myelomeningocele risk and ultra-rare deleterious variants in genes associated with cilium, WNT-signaling, ECM, cytoskeleton and cell migration. Sci Rep. 02 11 2021;11(1):3639. doi:10.1038/s41598-021-83058-7

54. Yu J, Wu Y, Yang P. High glucose-induced oxidative stress represses sirtuin deacetylase expression and increases histone acetylation leading to neural tube defects. J Neurochem. May 2016;137(3):371–83. doi:10.1111/jnc.13587

55. Taibi A, Mandavawala KP, Noel J, et al. Zebrafish churchill regulates developmental gene expression and cell migration. Dev Dyn. Jun 2013;242(6):614–21. doi:10.1002/dvdy.23958

56. Snir M, Ofir R, Elias S, Frank D. Xenopus laevis POU91 protein, an Oct3/4 homologue, regulates competence transitions from mesoderm to neural cell fates. EMBO J. Aug 09 2006;25(15):3664–74. doi:10.1038/sj.emboj.7601238

57. Zhang T, Lou J, Zhong R, et al. Genetic variants in the folate pathway and the risk of neural tube defects: a meta-analysis of the published literature. PLoS One. 2013;8(4):e59570. doi:10.1371/journal.pone.0059570

58. Griswold AJ, Ma D, Sacharow SJ, et al. A de novo 1.5 Mb microdeletion on chromosome 14q23.2-23.3 in a patient with autism and spherocytosis. Autism Res. Jun 2011;4(3):221–7. doi:10.1002/aur.186

59. Vannucchi H, Moreno FS. Interaction of niacin and zinc metabolism in patients with alcoholic pellagra. Am J Clin Nutr. Aug 1989;50(2):364–9. doi:10.1093/ajcn/50.2.364

60. Vannucchi H, Kutnink MD, Sauberlich M, Howerde E. Interaction among niacin, vitamin B6 and zinc in rats receiving ethanol. Int J Vitam Nutr Res. 1986;56(4):355–62.

61. Davis AP, Grondin CJ, Johnson RJ, et al. Comparative Toxicogenomics Database (CTD): update 2021. Nucleic Acids Res. 01 08 2021;49(D1):D1138–D1143. doi:10.1093/nar/gkaa891

62. Van Hateren N, Shenton T, Borycki AG. Expression of avian C-terminal binding proteins (Ctbp1 and Ctbp2) during embryonic development. Dev Dyn. Feb 2006;235(2):490–5. doi:10.1002/dvdy.20612

63. Aranow C. Vitamin D and the immune system. J Investig Med. Aug 2011;59(6):881–6. doi:10.2310/JIM.0b013e31821b8755

64. Chen Y-H, Yu Z, Fu L, et al. Supplementation With Vitamin D3 During Pregnancy Protects Against Lipopolysaccharide-Induced Neural Tube Defects Through Improving Placental Folate Transportation. Toxicological Sciences. 2015;145(1):90–97. doi:10.1093/toxsci/kfv036

65. Denny KJ, Jeanes A, Fathe K, Finnell RH, Taylor SM, Woodruff TM. Neural tube defects, folate, and immune modulation. Birth Defects Res A Clin Mol Teratol. Sep 2013;97(9):602–609. doi:10.1002/bdra.23177

66. Aguiar-Pulido V, Wolujewicz P, Martinez-Fundichely A, et al. Systems biology analysis of human genomes points to key pathways conferring spina bifida risk. Proc Natl Acad Sci U S A. 12 21 2021;118(51)doi:10.1073/pnas.2106844118

67. Kowitzke B, Cohrs G, Leuschner I, et al. Cellular Profiles and Molecular Mediators of Lesion Cascades in the Placode in Human Open Spinal Neural Tube Defects. J Neuropathol Exp Neurol. 09 2016;75(9):827–42. doi:10.1093/jnen/nlw057

68. Cohrs G, Blumenröther AK, Sürie JP, Synowitz M, Held-Feindt J, Knerlich-Lukoschus F. Fetal and Perinatal Expression Profiles of Proinflammatory Cytokines in the Neuroplacodes of Rats with Myelomeningoceles: A Contribution to the Understanding of Secondary Spinal Cord Injury in Open Spinal Dysraphism. J Neurotrauma. 12 2021;38(24):3376–3392. doi:10.1089/neu.2021.0091

69. Minihane AM, Vinoy S, Russell WR, et al. Low-grade inflammation, diet composition and health: current research evidence and its translation. Br J Nutr. Oct 14 2015;114(7):999–1012. doi:10.1017/S0007114515002093

70. Childs CE, Calder PC, Miles EA. Diet and Immune Function. Nutrients. Aug 16 2019;11(8)doi:10.3390/nu11081933

71. Weida J, Patil AS, Schubert FP, et al. Prevalence of maternal cell contamination in amniotic fluid samples. J Matern Fetal Neonatal Med. Sep 2017;30(17):2133–2137. doi:10.1080/14767058.2016.1240162

72. Sicherman J, Newton DF, Pavlidis P, Sibille E, Tripathy SJ. Estimating and Correcting for Off-Target Cellular Contamination in Brain Cell Type Specific RNA-Seq Data. Front Mol Neurosci. 2021;14:637143. doi:10.3389/fnmol.2021.637143

73. Blom HJ, Shaw GM, den Heijer M, Finnell RH. Neural tube defects and folate: case far from closed. Nat Rev Neurosci. Sep 2006;7(9):724–31. doi:10.1038/nrn1986

